# A Digital Therapeutic Intervention for Smoking Cessation in Adult Smokers: Randomized Controlled Trial

**DOI:** 10.1101/2020.06.25.20139741

**Authors:** Jamie Webb, Sarrah Peerbux, Peter Smittenaar, Sarim Siddiqui, Yusuf Sherwani, Maroof Ahmed, Hannah MacRae, Hannah Puri, Sangita Bhalla, Azeem Majeed

## Abstract

**Background:** Tobacco smoking remains the leading cause of preventable death and disease worldwide. Digital interventions delivered through smartphones offer a promising alternative to traditional methods, but little is known about their effectiveness.

**Objective:** Our objective was to test the effectiveness of Quit Genius, a novel digital therapeutic intervention for smoking cessation.

**Methods:** A two-arm, single-blinded, parallel-group randomized controlled trial design was used. Participants were recruited via referrals from primary care practices and social media advertisements in the UK. 556 Adult smokers (aged ≥18 years), smoking at least five cigarettes a day for the past year were recruited. 530 were included for the final analysis. Participants were randomized to one of two interventions. Treatment consisted of a digital therapeutic intervention for smoking cessation consisting of a smartphone application delivering cognitive behavioral therapy content, one-to-one coaching, craving tools and tracking capabilities. The control intervention was Very Brief Advice along the Ask, Advise, Act model. All participants were offered nicotine replacement therapy for three months. A random half of each arm was assigned a carbon monoxide (CO) device for biochemical verification. Outcomes were self-reported via phone or online. The primary outcome was self-reported 7-day point prevalence abstinence at 4-weeks post quit date.

**Results:** 556 participants were randomized (treatment n=277, control n=279). The intention-to-treat analysis included 530 participants (n=265 in each arm; 11 excluded for randomization before trial registration, and 15 for protocol violations at baseline visit). By the quit date (an average 16 days after randomization) 89% (236/265) of those in the treatment arm were still actively engaged. At the time of primary outcome, 74% (196) of participants were still engaging with the app. At 4-weeks post-quit date, 45% (118) of participants in the treatment arm had not smoked in the preceding 7-days, compared to 29% (76) in control (risk ratio 1.55, 1.23-1.96, *P* = .0002; intention-to-treat, N=530). Self-reported 7-day abstinence agreed with CO measurement (CO <10 ppm) in 96% of cases (80/83) where CO readings were available. No harmful effects of the intervention were observed.

**Conclusions:** The Quit Genius digital therapeutic intervention is a superior treatment in achieving smoking cessation four weeks post quit date compared to very brief advice.

**Trial Registration:** The trial was registered in the ISRCTN database on December 18, 2018 (https://www.isrctn.com/ISRCTN65853476).

## Introduction

Smoking is a leading cause of preventable and premature death worldwide. It is an important risk factor for serious health problems and life-threatening diseases [1]. Globally, tobacco use causes more than 8 million yearly deaths [2]. Moreover, the total global economic cost of smoking is more than $1.4 trillion a year [3]. Smoking is, therefore, a major worldwide economic and public health concern [1-3].

In the United Kingdom, Very Brief Advice (VBA) is the recommended clinical practice on smoking cessation for all health care practitioners [4-5]. It is designed to promote quit attempts and to be used opportunistically, in virtually any situation [4-5]. Those interested in quitting are referred to their local Stop Smoking Services (SSS) [4]. SSS typically combines face-to-face behavioral support with the option of pharmacotherapy, offered as nicotine replacement therapy (NRT) or varenicline. Similarly, in the United States, smokers access free telephone services and/or tobacco cessation websites that provide access to pharmacotherapy and additional behavioral support [6].

Despite traditional smoking cessation programs demonstrating efficacy, they only help ~15% of smokers to quit long-term [7]. Such programs have been shown to have limited utilization, due to scheduling, time and financial constraints [8-9]. Telephone support can overcome these barriers, but reaches only ~1% of smokers annually [10]. Given that support may be difficult to access, there is an urgent need for alternative solutions that are cost-effective, convenient and scalable.

Technological advancements have led to new approaches that aim to overcome the drawbacks of conventional smoking cessation programs. Digital health apps are one new approach with the potential to support behavior change [11-12]. They have been used successfully across a multitude of therapeutic areas including chronic conditions [13-17] and to promote healthy behaviors [18-23].

Smartphone apps have advantages over traditional approaches including ease of accessibility, personalization of interactions with real-time feedback, scalability to large populations, and cost-effectiveness [24]. In 2018, the number of mobile phone subscriptions topped eight billion globally [24]. Smartphone apps have the potential to reach smokers who would not, or who are unable to, utilize traditional services.

Yet, there is a paucity of data that examines the efficacy of smartphone apps for smoking cessation. A review of mobile phone-based smoking support identified only five randomized controlled trials (RCT) that tested the effectiveness of smartphone apps with low-intensity support, with each showing limited efficacy [24]. Furthermore, a recent content analysis revealed low adherence of existing smartphone apps to evidence-based treatment guidelines [25], whilst a review of the 50 most downloaded cessation apps found only two with scientific support [26].

Digital therapeutics are a new wave of digital health apps that can deliver high-intensity evidence-based therapeutic programs. Emerging evidence is encouraging and suggests that high-intensity support delivered via smartphone apps can aid smoking cessation. Yet, many early studies suffer from limitations such as utilizing single-arm cohorts or using non-CO verified self-reported abstinence to assess intervention efficacy [21-23]. To date, no RCT has been conducted on a high-intensity digital therapeutic intervention (DTI) for smoking cessation.

### Objectives

This study had several objectives. The first objective of this study was to test the effectiveness of the DTI Quit Genius (QG) by measuring 7-day point prevalence abstinence at 4-weeks post quit date. Other objectives included assessing user engagement with QG, as well as testing its effect on cognitive, attitudinal, and emotional outcomes.

## Methods

### Design

We conducted a two-arm, single-blinded, parallel-group, pre-registered randomized controlled trial, with 4-weeks, 6-months and 12-months follow-up. Here we report the primary outcomes at 4-weeks only as data collection for later time-points is still ongoing. Approval was granted by the Health and Social Care Research Ethics Committee A (HSC REC A; reference 18/NI/0171) and this research complies with the Declaration of Helsinki.

### Participants

Participants were recruited in the United Kingdom between January and November 2019. Adult smokers (aged ≥18 years) were invited to participate if they had been smoking at least 5 cigarettes a day for the past year, were not using any other form of stop smoking support, and had sufficient mobile phone functionality (5^th^ generation or higher for Apple iPhone or version 18 or higher for Android). The exclusion criteria included not speaking English, pregnancy, COPD, psychiatric medication, and a serious health condition that would substantially hinder completion of the intervention or control as determined by the study team. Participants with serious health conditions and/or using psychiatric medication were ruled out as a safety consideration.

Participants were recruited offline from primary care practices across London via SMS campaigns. Posters and leaflets at local community venues and advertisements on social media were also used. Recruitment advertisements and study information given to participants described the opportunity of being allocated to one of two possible behavioral interventions in addition to optional NRT. No mention of a digital intervention was made to the participants before randomization. Participants randomized onto the study received £10 to offset travel expenses. Participants completed a questionnaire online or via telephone at 4-weeks after their quit-date and were paid £20 for completion.

### Registration

The trial was registered in the ISRCTN database on December 18, 2018 (https://www.isrctn.com/ISRCTN65853476). On July 24, 2019, we adjusted the primary outcome to equate it with the majority of trials on smoking cessation. Specifically, the time window for assessing whether the participant had successfully quit was reduced from two weeks to one week. The previously approved primary outcome related to the Russell Standard (i.e. self-reported abstinence in the past 2-weeks at 4-weeks post quit date). This was changed to capture self-reported 7-day point prevalence abstinence at 4-weeks. This decision was informed in-part by a recent Delphi study which found only partial compliance with the Russell Standard, as reported by smoking cessation experts [27]. Although we acknowledged a lack of consensus relating to outcome criteria in smoking cessation research, it was of our opinion that the use of 7-day point prevalence is preferable to 14-day point prevalence as it allows for greater comparability to other studies. Amending the primary outcome to 7-day point-prevalence allowed for greater comparison with other studies of face-to-face, digital and other low intensity intervention smoking cessation trials(full justification is available on the ISRCTN page). This decision was made without having analyzed trial outcomes. The 14-day point prevalence was added as a secondary outcome. Another secondary outcome (“number of quit attempts up to week four post-quit date”) is reported here as “Any additional quit attempt after the quit date” because a continuous test was not appropriate on the distribution which was predominantly one quit attempt. All other adjustments to the trial registration only concerned start/end dates of recruitment and publication to account for unexpected challenges in recruitment.

### Randomization and masking

Participants were randomized 1:1 (treatment:control) using a block size of four participants through a trial management system (www.curebase.com). Researchers randomizing participants were blind to allocation until they had performed the randomization.

### Procedures

At first contact (via phone or online), participants were provided with study information and completed a questionnaire to determine eligibility. If eligible, participants were invited to attend an in-person baseline session where eligibility was reconfirmed and informed consent and baseline data was collected. Participants were then randomized. All participants were recommended to set their quit date within two-weeks of randomization, however this was not a mandatory requirement for study participation. All participants were offered NRT in addition to their allocated intervention. At 4-weeks post-quit-date participants were invited to complete the follow-up survey (via phone or online).

### Treatment intervention: Quit Genius

Quit Genius (QG) is a smartphone application comprising a comprehensive digital solution informed by the principles of cognitive behavior therapy (CBT) (Screenshots shown in Figure 1). It is a year-long program designed to support the user both before and after their quit date. Quit Genius delivers intervention components that have demonstrated efficacy in promoting smoking cessation, including self-monitoring, goal setting, encouraging medication adherence, and feedback on progress (www.quitgenius.com). QG was developed over many iterations including engagement with smokers, patient representatives and scientific advisors. The application collects data on users through in-app metrics to help personalize the program. Metrics include usage, session completion, program completion and quit date. Additional data are collected based on user participation and feedback following CBT exercises, giving information such as their reasons for quitting smoking and the reasons why they continue to smoke.

**Figure 1:**
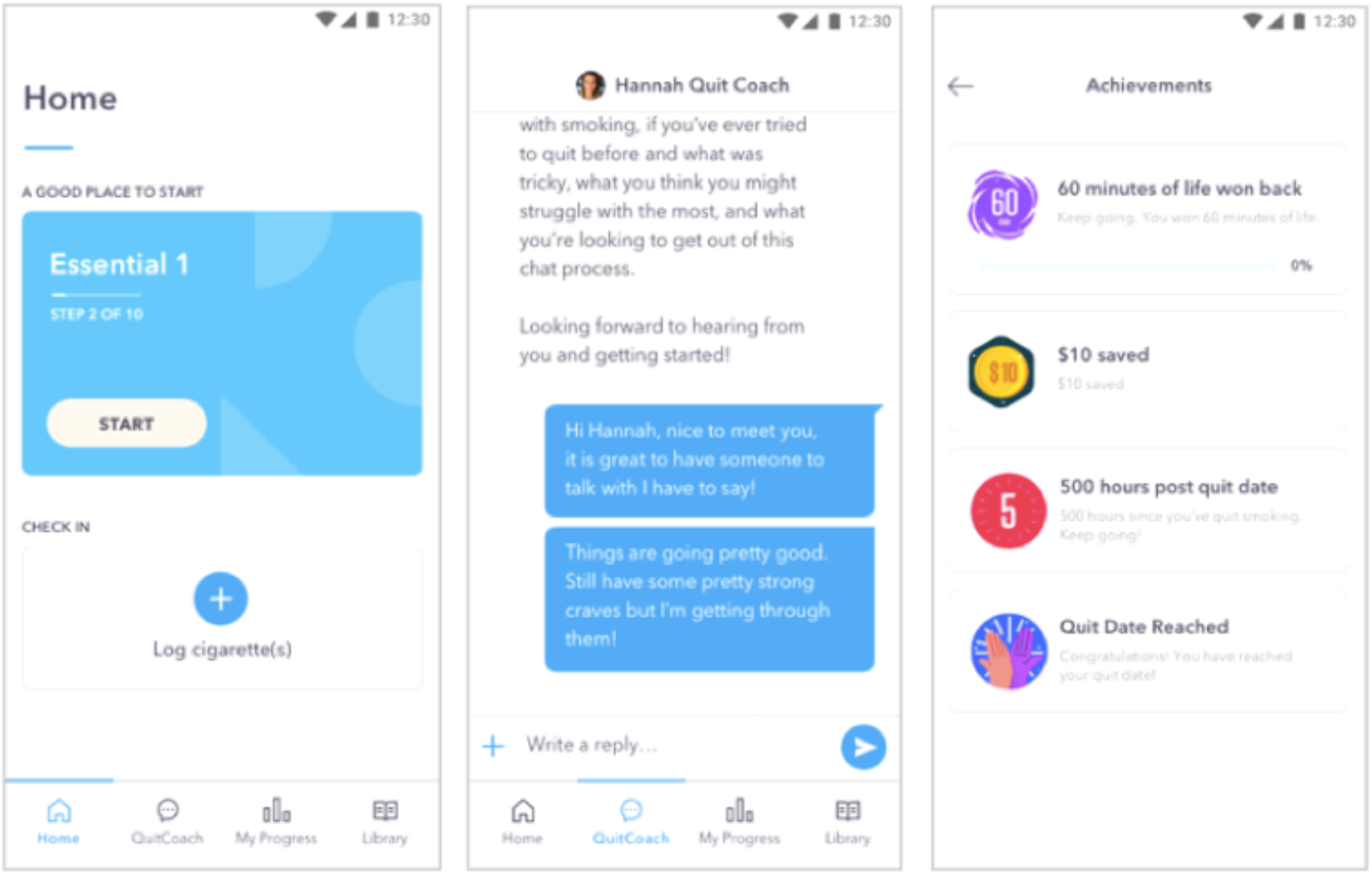
Screenshots of the digital therapeutic intervention Quit Genius.

Content is tailored to the user and is delivered in the form of animated videos, audio sessions, reflective exercises, and quizzes. The user is prompted to complete a series of self-paced steps on their smoking cessation journey, with each new step (and content) unlocking only once the previous step has been completed. The program content is divided into the ‘Essentials’ stage, which the user is prompted to complete before their quit-date, and covers aspects such as preparing for the quit date, using nicotine replacement therapy, and thinking about the reasons for quitting. The ‘Sustain’ stage, which the user completes once they quit smoking, focuses on the general principles of relapse prevention and helps the user to stay smoke-free in the long-term. In the time leading up to their quit date, users are encouraged to monitor their smoking habits daily by logging the number of cigarettes smoked, their triggers (how they felt when they wanted to smoke), and the intensity of their craving. Once the user has quit smoking, they are encouraged to log whether or not they are currently smoking.

As part of the QG program participants also have access to a ‘Quit Coach’, an advisor qualified by the National Centre for Smoking Cessation and Training (NCSCT). The coach provides personalized support via a digital chat interface and phone. Typically, participants partake in one initial phone call with the rest of quit coach interaction mediated through the in-app digital chat interface. Users can monitor their progress via the app, which details improvements to health and any financial benefits gained from being a non-smoker since their quit date. Finally, users can access a ‘Craving Toolbox’ which comprises audio content of short breathing exercises, mindfulness exercises and meditation exercises designed to help the user manage their cravings to smoke. The QG app uses CBT to target not only smoking cessation but also covers skills and strategies to promote improved mental health and wellbeing. Whilst the QG app does not substitute professional care for mental health concerns, the app specifically addresses common mental health concerns such as low mood, anxiety, stress, self-esteem, and social skills. The app also specifically targets general health and wellbeing concerns such as diet, exercise, and self-care techniques. Specific skills and techniques used include goal setting, cognitive restructuring, graded exposure, progressive muscle relaxation, mindfulness, assertiveness and communication training, and problem-solving skills. QG users receive push notifications to serve as reminders to engage with the app. All participants in the treatment group received free access to the QG intervention.

### Control group: Very Brief Advice

VBA is a simple form of advice designed to be used opportunistically. It follows the Ask, Advise, Act structure as recommended by the UK government. Participants were advised to contact their local stop smoking service to access support and medication to quit smoking. Trial assistants were trained in the delivery of VBA as per NCSCT guidelines (https://elearning.ncsct.co.uk). Of the control group participants allocated a CO device, a non-branded mobile-app (ASH-app) was also provided to visualize CO readings for the participant. The control group mobile-app was only used in conjunction with the CO device and contained no other content for participants.

### Nicotine Replacement Therapy

All participants had the option to receive nicotine replacement products (2mg or 4mg gum and/or 16hr or 24hr patches) free for 12 weeks, with the first two-week supply issued at the baseline visit. Participants were allowed to purchase alternative forms of oral NRT.

### Carbon Monoxide Monitor

Half of all participants were given a carbon monoxide (CO) monitor to measure levels of carbon monoxide in their breath and to validate self-reported smoking abstinence. Participants were selected pseudorandomly to ensure 50% of each group was assigned a device. CO levels were collected via self-report at 4-weeks post quit-date. The CO devices plugged into the headphone/charging slot of participants’ smartphones, and were used in conjunction with the QG and control app. At the follow-up time-point, participants were asked to give a reading from their device via phone or online. NCSCT guidelines of a CO reading <10ppm were used to validate participants’ self-reported abstinence [28].

### Measures

Measurements were taken at baseline and at the 4-week follow-up. The following variables were collected at baseline: demographic details, smoking status, smoking history, expired carbon monoxide level, Fagerstrom Test for Nicotine Dependence (FTND) [29], Smoking Abstinence Self-efficacy Questionnaire (SASEQ) [30], Warwick-Edinburgh Mental Well-being Scale (WEMWBS) [31], WHOQOL-BREF [32] and the Service Use Questionnaire (SUQ) [33]. At 4-weeks post quit-date, smoking status, changes in attitudes and perceptions of smoking, SASEQ, WEMWBS, and WHOQOL-BREF were collected. Expired carbon monoxide level was collected only in those participants who were assigned their own device (Smokerlyzer, coVita Inc.). Measurements were collected via online questionnaires.

The primary outcome was self-reported 7-day point prevalence abstinence at 4-weeks post quit-date. Secondary outcomes at week four included 14-day point prevalence abstinence, any additional quit attempts after the quit-date, self-reported changes in confidence levels, knowledge, attitudes and perceptions related to smoking cessation, changes in SASEQ and WEMWBS, and satisfaction with the treatment intervention (treatment group only).

### Data Analysis

At 4-weeks, we expected to observe a 7-day abstinence rate of 25% in the treatment group and 10% in the control group. At a Type I error rate of 5% and power of 90% we required 133 participants per group (266 total). At 6-months, a conservative estimate would be a 10% quit rate in the treatment group and 3% in the control group. To detect a difference with 80% power and 5% Type I error we needed to randomize 194 participants per group (388 total). Assuming a 20% dropout, we aimed to recruit ≥500 participants.

We performed both intention-to-treat (ITT) [34] and per-protocol (PP) analyses. ITT included all participants assigned to treatment and control, respectively, as indicated in Figure 1. ITT analysis assumed that participant data was not missing at random. PP included the subset of participants that provided answers to the self-reported outcomes at week-4.

We used chi-squared tests for binary outcomes and two-sample t-tests for continuous outcomes. Those lost to follow-up at 4-weeks were considered “currently smoking” for the primary outcome as well as 2-week abstinence; no additional quit attempts after the quit date; not “strongly agree” to improvements in confidence, knowledge, or attitude; and no change in self-efficacy or mental well-being from baseline. We used logistic regression to estimate the main effect of being assigned a CO device on likelihood of quitting, with treatment assignment as a covariate in the model.

All data processing and analysis was performed in R using the *tidyverse* package family and the *fmsb* package [35–37].

## Results

### Participant flow chart

Figure 2 shows the CONSORT flowchart for the RCT.

**Figure 2:**
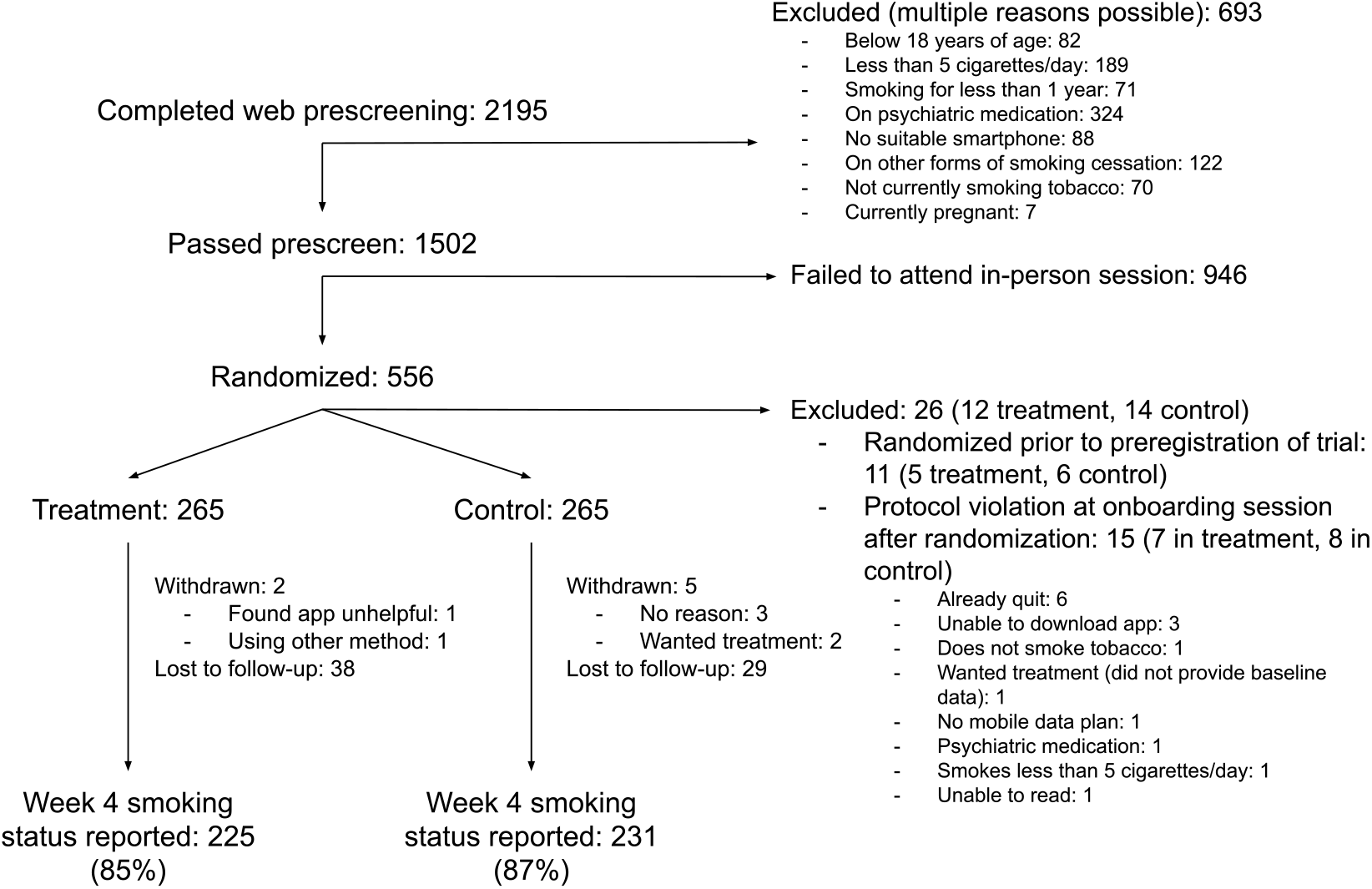
CONSORT flow chart.

### Participants

556 participants were randomized (treatment n=277, control n=279). The intention-to-treat analysis included 530 participants (n=265 in each arm; 11 excluded for randomization before trial registration, and 15 for protocol violations at baseline visit). Participants came from a wide age range, were slightly more likely to be male than female, and about two in three self-identified as Caucasian/White (Table 1). Educational attainment ranged uniformly from secondary education up to postgraduate education, and 80.0% (424/530) were in paid employment, of which over half were in “Managerial or professional” roles. Participants were smoking on average 14 (treatment) or 15 (control) cigarettes per day, with nicotine dependence of four out of 10 on the Fagerstrom Test for Nicotine Dependence. Most participants (85.0%, 451/530) had previously made quit attempts, primarily going cold turkey or with the help of e-cigarettes and NRT. No substantial differences between treatment and control groups were introduced through randomization.

**Table 1:** Demographics and smoking history of treatment and control groups. SD: standard deviation. NRT: nicotine replacement therapy.

|  | Treatment | Control |
| --- | --- | --- |
| Number of participants | 265 | 265 |
| **Age (SD)** | 40 (12) | 42 (12) |
| **Female** | 46.0% | 44.0% |
| **Ethnicity** |  |  |
| Caucasian/White | 69.0% | 62.0% |
| Black/Caribbean/African/Black | 9.0% | 11.0% |
| Asian | 7.0% | 9.0% |
| Arab | 1.0% | 2.0% |
| Mixed | 9.0% | 9.0% |
| Other | 3.0% | 4.0% |
| Prefer not to say | 2.0% | 3.0% |
| **Education** |  |  |
| GCSE or lower | 22.0% | 23.0% |
| A-level | 25.0% | 19.0% |
| Undergraduate degree | 29.0% | 29.0% |
| Postgraduate degree | 17.0% | 19.0% |
| PhD | 2.0% | 1.0% |
| Prefer not to say | 6.0% | 9.0% |
| **In paid employment** | 79.0% | 81.0% |
| **Type of employment (if employed)** |  |  |
| Managerial or professional | 60.0% | 53.0% |
| Routine or manual | 10.0% | 15.0% |
| Intermediate | 10.0% | 9.0% |
| Other | 18.0% | 19.0% |
| Prefer not to say | 1.0% | 3.0% |
| **Cigarettes per day (SD)** | 14 (6) | 15 (7) |
| **Fagerström Test for Nicotine Dependence (range 0-10; SD)** | 4 (2) | 4 (2) |
| **Any past attempt to quit smoking** | 84.0% | 86.0% |
| **Method previously used (if past attempts)** |  |  |
| Cold turkey | 47.0% | 49.0% |
| E-cigarettes | 42.0% | 42.0% |
| NRT | 31.0% | 28.0% |
| Prescription medication | 11.0% | 15.0% |
| Smartphone app | 9.0% | 10.0% |
| Hypnotherapy | 4.0% | 7.0% |
| Psychological Therapy | 2.0% | 2.0% |

### Engagement

Engagement with different facets of the DTI is shown in Table 2. By the quit date (an average 16 days after randomization) 89.0% (236/265) of those in the treatment arm were still actively engaged. At the time of primary outcome, 74.0% (196) of participants were still engaging with the app. The content consisting of education and cognitive behavioral therapy (“Essentials” 1 and 2) were consumed by 55.0% (146) and 36% (95) of participants, respectively. 69.0% (183) sent at least one in-app message to their coach, and on average people messaged their coach about once per week. On average, participants reported 12 diary entries to report cigarettes smoked before their quit date, and six check-ins to report cravings or lapses after their quit date.

**Table 2:** Engagement with the digital therapeutic program in the treatment group (intention-to-treat, n = 265). Essentials 1: program content aimed at preparation for the quit date. Essentials 2: program content intended for just after the quit date. Diary entry: registration of a cigarette smoked prior to quit date. Check-in: registration of a cigarette after the quit date.

| **Engagement** | **Value (mean (SD), 25/75 percentile)** |
| --- | --- |
| App opens up to week four | 37 (52), 9-43 |
| Days between randomization and quit date | 16 (21), 9-23 |
| Still active by quit date | 89% |
| Active four weeks after quit date | 74% |
| Completed Essentials 1 | 55% |
| Completed Essentials 2 | 36% |
| Sent 1+ messages to coach | 69% |
| Messages to coach before quit date | 3.3 (5.9), 0-4 |
| Messages to coach from quit date to week four | 4.2 (7.5), 0-6 |
| Messages from coach before quit date | 6.6 (6.3), 2-9 |
| Messages from coach from quit date to week four | 6.2 (5.8), 2-9 |
| Number of diary entries before quit date | 12 (27), 1-12 |
| Number of check-ins from quit date to week four | 6 (10), 0-9 |

### Outcomes

Table 3 shows the primary outcome and secondary outcomes for intention-to-treat and per-protocol analyses. In the intention-to-treat analysis, those in the treatment arm were 55% more likely to report 7-day abstinence 4-weeks after their quit date compared to those in control (RR: 1.55, 95% CI 1.23-1.96; 45% versus 29% quit rate). In those participants that were assigned a CO device (138/265 in treatment, 142/265 in control), 97% (134) in treatment and 98% (139) in control provided a CO reading at baseline, and 61% (82) and 67% (95) respectively provided a reading at 4-weeks (including those that did not complete the week-4 questionnaire). CO completion was 88% (50/57) and 89% (33/37) respectively in participants that claimed abstinence at 4-weeks. For these abstaining participants the CO measurement was below 10 parts per million (ppm) for 96% (48/50) and 97% (32/33) of participants in treatment and control, respectively. Whether or not a participant was provided with a CO device did not significantly predict quit rate (p=.29 in logistic regression with CO device and intervention main effects). There was no difference in rate of NRT use in treatment and control (59.0% and 63.0% respectively in those that completed week-4 questionnaire; RR: 0.94, 95% CI 0.81-1.08)

**Table 3:**
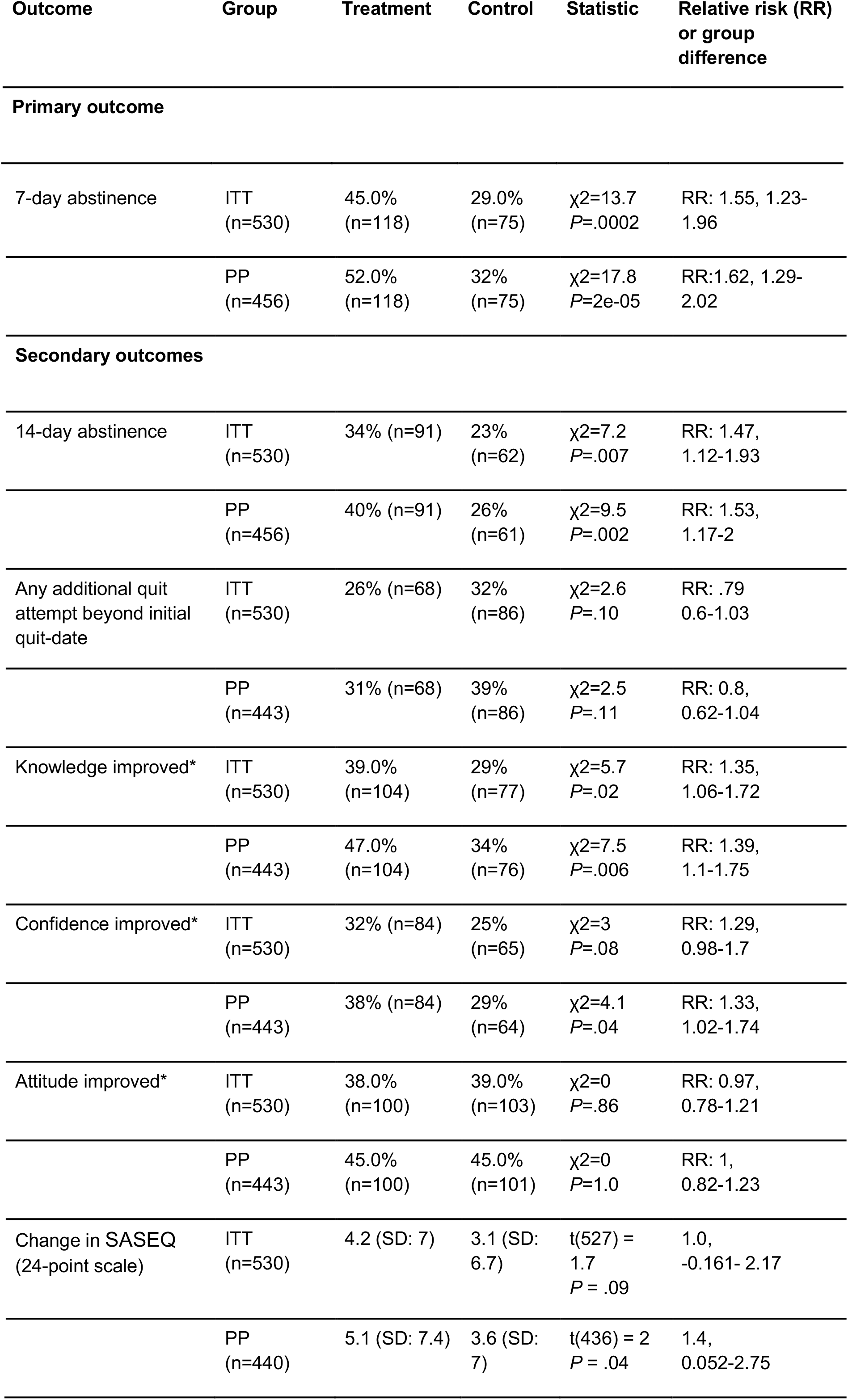
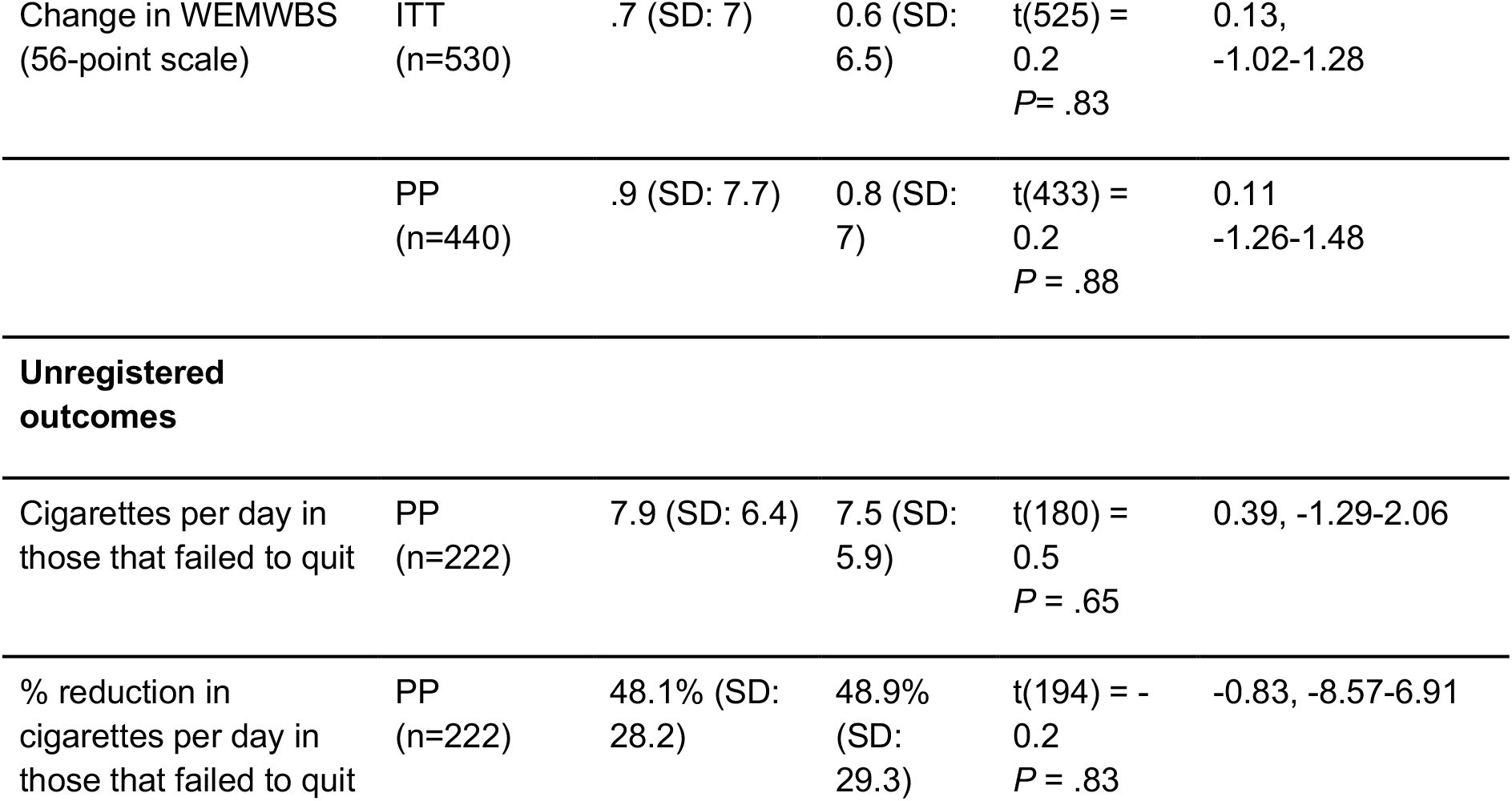
Outcomes at four weeks after quit date. ITT: intention-to-treat. PP: per protocol. WEMWBS: Warwick-Edinburgh Mental Wellbeing Scale. SASEQ: Smoking abstinence self-efficacy questionnaire. * as measured by % of participants reporting “strongly agree”.

| Outcome | Group | Treatment | Control | Statistic | Relative risk (RR) or group difference |
| --- | --- | --- | --- | --- | --- |
| **Primary outcome** |  |  |  |  |  |
| 7-day abstinence | ITT<br>(n=530) | 45.0%<br>(n=118) | 29.0%<br>(n=75) | $\chi^2=13.7$<br>$P=.0002$ | RR: 1.55, 1.23-1.96 |
| | PP<br>(n=456) | 52.0%<br>(n=118) | 32%<br>(n=75) | $\chi^2=17.8$<br>$P=2e-05$ | RR: 1.62, 1.29-2.02 |
| **Secondary outcomes** |  |  |  |  |  |
| 14-day abstinence | ITT<br>(n=530) | 34% (n=91) | 23%<br>(n=62) | $\chi^2=7.2$<br>$P=.007$ | RR: 1.47,<br>1.12-1.93 |
| | PP<br>(n=456) | 40% (n=91) | 26%<br>(n=61) | $\chi^2=9.5$<br>$P=.002$ | RR: 1.53,<br>1.17-2 |
| Any additional quit attempt beyond initial quit-date | ITT<br>(n=530) | 26% (n=68) | 32%<br>(n=86) | $\chi^2=2.6$<br>$P=.10$ | RR: .79<br>0.6-1.03 |
| | PP<br>(n=443) | 31% (n=68) | 39%<br>(n=86) | $\chi^2=2.5$<br>$P=.11$ | RR: 0.8,<br>0.62-1.04 |
| Knowledge improved* | ITT<br>(n=530) | 39.0%<br>(n=104) | 29%<br>(n=77) | $\chi^2=5.7$<br>$P=.02$ | RR: 1.35,<br>1.06-1.72 |
| | PP<br>(n=443) | 47.0%<br>(n=104) | 34%<br>(n=76) | $\chi^2=7.5$<br>$P=.006$ | RR: 1.39,<br>1.1-1.75 |
| Confidence improved* | ITT<br>(n=530) | 32% (n=84) | 25%<br>(n=65) | $\chi^2=3$<br>$P=.08$ | RR: 1.29,<br>0.98-1.7 |
| | PP<br>(n=443) | 38% (n=84) | 29%<br>(n=64) | $\chi^2=4.1$<br>$P=.04$ | RR: 1.33,<br>1.02-1.74 |
| Attitude improved* | ITT<br>(n=530) | 38.0%<br>(n=100) | 39.0%<br>(n=103) | $\chi^2=0$<br>$P=.86$ | RR: 0.97,<br>0.78-1.21 |
| | PP<br>(n=443) | 45.0%<br>(n=100) | 45.0%<br>(n=101) | $\chi^2=0$<br>$P=1.0$ | RR: 1,<br>0.82-1.23 |
| Change in SASEQ (24-point scale) | ITT<br>(n=530) | 4.2 (SD: 7) | 3.1 (SD: 6.7) | $t(527) = 1.7$<br>$P = .09$ | 1.0,<br>-0.161- 2.17 |
| | PP<br>(n=440) | 5.1 (SD: 7.4) | 3.6 (SD: 7) | $t(436) = 2$<br>$P = .04$ | 1.4,<br>0.052-2.75 |
| Change in WEMWBS<br>(56-point scale) | ITT<br>(n=530) | .7 (SD: 7) | 0.6 (SD:<br>6.5) | t(525) =<br>0.2<br>P= .83 | 0.13,<br>-1.02-1.28 |
|  | PP<br>(n=440) | .9 (SD: 7.7) | 0.8 (SD:<br>7) | t(433) =<br>0.2<br>P = .88 | 0.11<br>-1.26-1.48 |
| **Unregistered**<br>**outcomes** |  |  |  |  |  |
| Cigarettes per day in<br>those that failed to quit | PP<br>(n=222) | 7.9 (SD: 6.4) | 7.5 (SD:<br>5.9) | t(180) =<br>0.5<br>P = .65 | 0.39, -1.29-2.06 |
| % reduction in<br>cigarettes per day in<br>those that failed to quit | PP<br>(n=222) | 48.1% (SD:<br>28.2) | 48.9%<br>(SD:<br>29.3) | t(194) = -<br>0.2<br>P = .83 | -0.83, -8.57-6.91 |

For secondary outcomes, 14-day abstinence showed greater efficacy of treatment compared to control (RR: 1.47, 95% CI 1.12-1.93). Those in treatment were no more or less likely to have made an additional quit attempt after the initial quit date (RR: 0.79, 95% CI 0.60-1.03). Those in the treatment arm were more likely to “strongly agree” that their knowledge of their smoking habit had improved (RR: 1.35, 95% CI 1.06-1.72), but no such effect was observed regarding their confidence in their ability to stay smoke-free (RR: 1.29, 95% CI 0.98-1.70]) nor in terms of whether their attitude towards stopping smoking had become more positive (RR: 0.97, 95% CI 0.78-1.21). The treatment was also not superior to control in terms of the increase in smoking self-efficacy (*P* = .09), nor in terms of mental well-being (P = .83). However, in the per-protocol analysis, which included only participants that completed their week-4 outcomes, several secondary outcomes were significantly better in treatment compared to control: confidence improved more (RR: 1.33, 95% CI 1.02-1.74), as did self-efficacy (P = .04).

Participant satisfaction with the treatment intervention was high (209/265 in the treatment group that completed the questionnaire at week four). On a scale from zero (least satisfied) to three (most satisfied): quality of the smoking cessation service: 2.4, SD 0.7; program meeting needs: 2.2, SD 0.8; helpfulness of information: 2.6, SD 0.6; helped to deal with smoking effectively: 2.4, SD 0.7; likelihood of coming back if needing to quit in the future: 2.5, SD 0.7. Asked whether the participant would recommend the digital therapeutic to a friend, 92% (192) would do so. Participants considered the quit coach and the education/CBT articles to be most helpful, whereas the community element of the program was considered least helpful (as shown in Figure 3).

**Figure 3:**
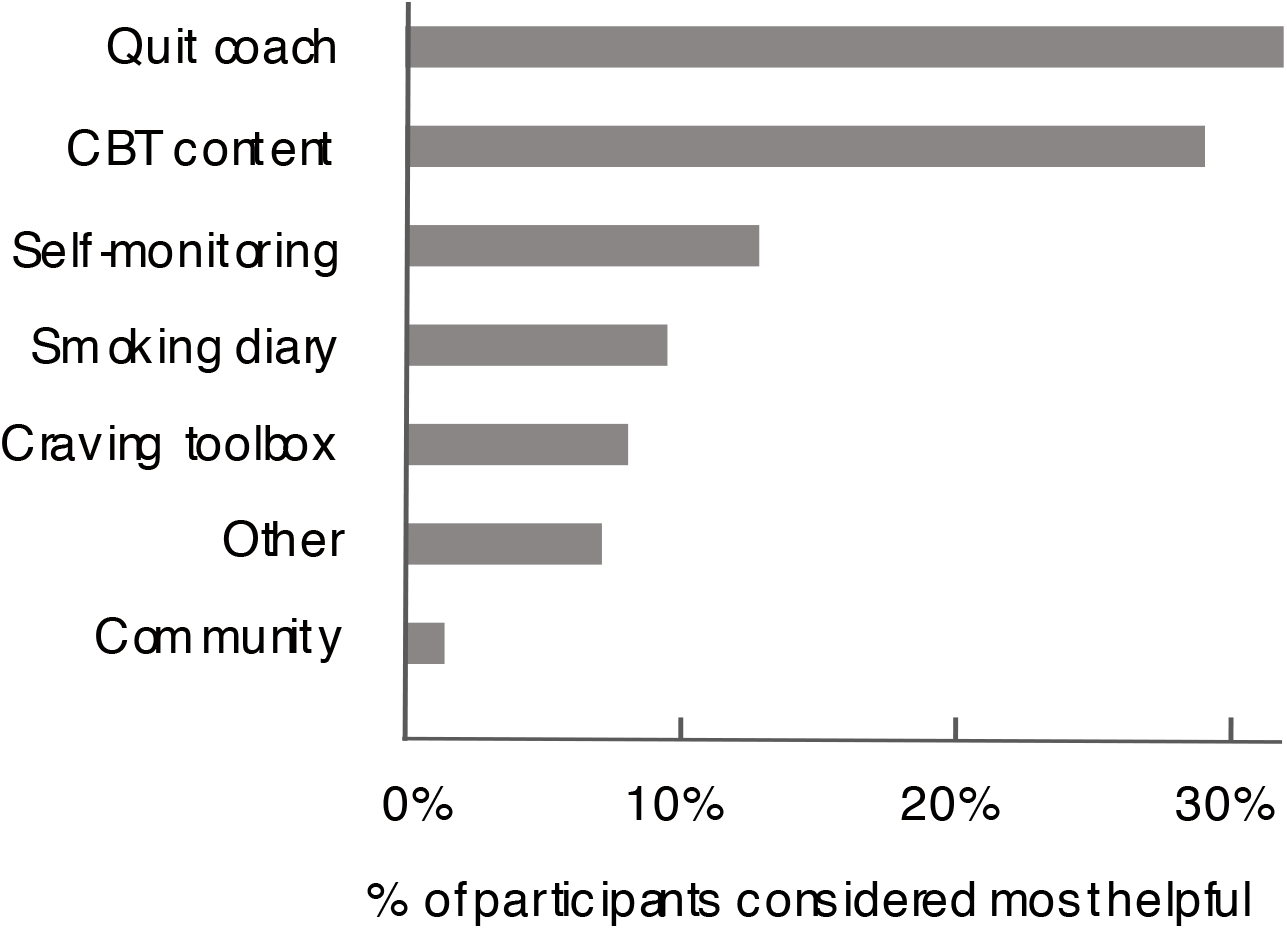
Elements of the program considered most helpful by n=209 participants who completed the user satisfaction questionnaire at week four.

Finally, we examined two outcomes unregistered at trial registration. First, cigarettes smoked per day by those that failed to quit showed no difference between treatment and control (treatment: 7.9, control: 7.5 cigarettes per day). Similarly, the average % reduction in cigarettes per day, though substantial in both groups, showed no difference between groups (48.1% reduction in treatment vs 48.9% in control; *P* = .83).

## Discussion

### Principal findings

In this RCT we assessed the efficacy of Quit Genius, a comprehensive DTI for smoking cessation. The primary outcome of self-reported 7-day abstinence at 4-weeks post quit date was significantly higher for the treatment group compared to a control group using VBA.

Principally, this shows the superiority of the DTI compared to the UK’s typical first-line intervention for smoking cessation [4-5]. We show that a digital, remote-first program is an effective method for short-term behavior change. The treatment group demonstrated a CO-verified 4-week quit rate comparable with high-intensity face-to-face smoking cessation programs utilized by healthcare services in the United Kingdom [7]. Our results support previous literature illustrating that high-intensity behavioral support combined with pharmacotherapy is an effective means of quitting smoking [7].

Compared with other digital interventions, the 45.0% (ITT) and 52.0% (PP) seven-day abstinence rates four weeks after the quit date compare favorably to previous digital intervention studies. Pivot, another digital program, achieved a CO-verified benchmark seven-day abstinence rate of 32% (ITT) and 37% (PP) [21]. An Acceptance and Commitment Therapy (ACT) intervention by Smartquit achieved 21% 7-day abstinence [22]. Clickotine’s 7-day abstinence rate of 45% was similar to that observed in the current study [23]. However, all used single-arm designs rather than RCTs, and were therefore unable to distinguish between the causal effect of treatment, placebo, or underlying differences in propensity to quit in the study population [38]. Finally, both Smartquit and Clickotine studies used non-CO-verified self-reported abstinence as their primary means of assessing intervention efficacy, leaving a possibility of falsely reported smoking status [22-23]. This study avoided such limitations by using a two-arm, parallel-group RCT design with biochemical verification. Though short-term abstinence rates of the program studied here are promising both in absolute terms as well as compared to the control group, 6- and 12-month abstinence rates will confirm whether the program is efficacious in the longer term.

### Cognitive, attitudinal, and emotional improvements

Education and confidence are integral mechanisms in eliciting successful smoking cessation [39-40]. In this study, treatment caused a greater improvement in knowledge of personal smoking habits compared to control. Similarly, confidence in ability to stay smoke-free was higher in treatment than control, though only statistically significantly so in participants that completed the study per protocol. No effect was found on the attitude of participants towards stopping smoking. This suggests that the DTI is an effective tool for making people aware of their habit and instilling some degree of confidence, but fails to improve a commonly negative attitude towards stopping smoking. Given that smoking cessation interventions may be enhanced by incorporating strategies that target attitude change [41], the DTI could be further improved to engender a more positive attitude towards quitting.

Self-efficacy is a robust indicator of future successful smoking abstinence (40). We observed the DTI to be superior to control in improving self-efficacy in participants that completed the study per protocol, but not when analyzed by intention-to-treat. This resembles the increase in reported confidence, indicating that the DTI enhanced users’ beliefs in their capacity to quit successfully but that further developments on the DTI should focus on strengthening these outcomes.

We observed no benefit of the DTI compared to control in terms of mental well-being. In a retrospective study of the same DTI we observed a correlation between hedonic well-being and quit rates [42]. In line with this, smoking cessation is typically associated with improved mental health with evidence illustrating a reduction in anxiety, depression and stress after quitting [43]. Given the higher quit rates in the treatment group, we expected treatment to be superior in terms of mental well-being. However, improvements are usually demonstrated in longer-term follow-ups than the four weeks reported here [43]. As such, changes in mental well-being might not yet have manifested, and our 6-month and 12-month outcomes will shed light on the longer-term impact of the DTI on mental well-being. There are both positive and negative effects associated with smoking and smoking abstinence that could impact mental well-being. Evidence suggests that smokers who reduce their smoking but fail to quit show more pronounced mood deterioration than those who succeed [43]. Conversely, there is evidence illustrating the negative sequelae of smoking cessation, such as anxiety, insomnia and weight gain [44]. Therefore, it is possible that simultaneous effects could have been active within groups, resulting in no net effect on mental well-being.

Nicotine addiction is a condition that rarely exceeds a lifetime abstinence rate of >50% per-treatment. For this reason, many smokers take 30 or more quit attempts before being successful [45]. Nonetheless, a reduction in daily cigarette use predicts future behavior change: individuals who reduce their cigarette use by ~50% are more likely to see future quit attempts and greater odds of successful quitting [45]. Among treatment participants who had not quit smoking, there was an average reduction of 48% in their cigarette use compared to baseline, similar to control. Digital programs can, perhaps more easily than traditional programs, leverage such data to continue to engage and encourage specific participants after a failed quit attempt, identifying an optimal time to engage when chances of quitting are highest.

### Engagement

To elicit the success of any non-invasive behavioral intervention, users must actively partake in the treatment [17, 23]. We assessed engagement across several elements of the DTI, and observed 74% (196/265) were still using the app four weeks after their quit date. One differentiating aspect of this DTI from typical smartphone applications is the presence of human coaching. We found a consistent bidirectional flow of communication between quit coach and participant from baseline to quit date and from quit date to 4-week follow-up. Of all the elements of treatment offered by the DTI, coaching was considered most helpful. This reflects previous notions that smoking cessation programmes that utilize health coaching as a means of support are effective in eliciting successful smoking abstinence [46].

### Strengths and limitations

Particular strengths of this study are the randomized controlled design, preregistration of the trial and its outcomes, biochemical verification despite the remote nature of the program, and the high four-week follow-up rate. The key limitation of this study is the short follow-up period of 4-weeks. It is known that relapse occurs over a longer timeframe [47-49], and the current findings do not speak to the DTI’s ability to avoid longer-term relapse.

Another limitation is that participants may have exaggerated their self-reported smoking abstinence. To combat this, participants were informed from study outset that regardless of smoking abstinence, they could remain on the trial and would be eligible for remuneration. Another preventative measure was the use of a measurement device to assess CO levels in the exhaled breath of the individual. Due to cost considerations, these devices were provided to a random 50% of each study arm. The near-perfect agreement (80/83, 96%) between CO monitor and self-reported abstinence suggests that at least in the context of this DTI, self-report can be taken at face value. Lastly, the lack of experimenter blinding to participant group allocation may have introduced bias into data interpretation. To avoid this, standardized questionnaires, participant interaction scripts, and standard operating procedures were used across treatment groups, so that any effect on participant outcome data was minimized. Also, the trial outcome measures, and sample size were preregistered, and data was only analysed after data collection had been completed.

### Conclusions

A digital therapeutic intervention for smoking cessation was superior to very brief advice in achieving smoking cessation after 4-weeks in a biochemically verified RCT. The DTI examined here is an effective option for short-term smoking cessation. Participants were actively engaged with the DTI and were satisfied with the intervention. Nevertheless, opportunities exist to improve mental wellbeing and attitudinal outcomes. A critical open question pertains to the long-term efficacy, which will be reported in a subsequent paper.

## Data Availability

Data is available upon request

## Acknowledgements

The authors thank the participating NHS primary care practices located across the London districts of Barnet, Camden, Ealing, Hammersmith and Fulham, Islington, Kilburn and North Kensington for their support in participant recruitment. AM is supported by the NW London NIHR Applied Research Collaboration.

## Conflict of Interests

The study was funded by the company that produced the Quit Genius digital therapeutic intervention (Digital Therapeutics, Inc). PS is a paid statistical consultant. All other authors except AM received a salary from or own equity in Digital Therapeutics, Inc.

**Abbreviations**
ACT: acceptance and commitment therapy
CBT: cognitive behavioral therapy
CO: carbon monoxide
DTI: digital therapeutic intervention
ITT: intention-to-treat
NRT: nicotine replacement therapy
PP: per-protocol
QG: quit genius
RCT: randomized controlled trial
VBA: very brief advice

